# FOCUSED REVIEW ON NUTRITIONAL STATUS OF PATIENTS WITH IMMUNOGLOBULIN LIGHT CHAIN AMYLOIDOSIS

**DOI:** 10.1101/2021.04.14.21255509

**Authors:** Shana Souza Grigoletti, Priccila Zuchinali, Émilie Lemieux-Blanchard, Stéphanie Béchard, Bernard Lemieux, Paula Aver Bretanha Ribeiro, François Tournoux

## Abstract

**Background:** Immunoglobulin light chain (AL) amyloidosis is a complex disease marked by a poor clinical portrait and prognosis generally leading to organ dysfunction and shortened survival. We aimed to review the available evidence on whether AL amyloidosis can lead to malnutrition, thus having a negative impact on quality of life (QoL) and survival.

**Materials:** We searched Pubmed with no restrictions to the year of publication or language. Retrospective or prospective, observational, and interventional studies that reported data regarding AL amyloidosis and nutritional status were included.

**Results:** From 62 articles retrieved, 23 were included. Malnutrition was prevalent in up to 65% of patients with AL Amyloidosis. Prevalence of weight loss of 10% or more ranged from 6 to 22% of patients, while a body mass index of < 22 kg/m2 was found in 22 to 42%. Weight loss, lower BMI and other indicators of poor nutritional status were negatively associated with quality of life and survival. Only one RCT focused on nutritional counseling was found and reported positive results on patients QoL and survival.

**Conclusion:** Despite inconsistencies across assessment criteria, the available data reveal that weight loss and malnutrition are common features in patients with AL amyloidosis. This review reinforces the premise that an impaired nutritional status can be negatively associated with QoL and survival in patients with AL amyloidosis, and therefore should be further investigated.

## INTRODUCTION

Amyloidosis is a disorder in which insoluble amyloid fibrils accumulate in the extracellular space of organs and tissues, causing irreversible organ dysfunction and death ^1, 2^. Amyloidosis can be localized or systemic. Organs commonly involved include the kidneys, heart, peripheral and autonomic nervous system, liver, and gastrointestinal tract ^3, 4^.

There are several types of amyloidosis, one of the most common types in developed countries being immunoglobulin light chain (AL) amyloidosis ^3, 5^. The clinical portrait of AL amyloidosis varies substantially and depends largely on the disease stage and involved organ(s). Clinical manifestations such as restrictive cardiomyopathy, nephrotic syndrome, hepatic failure, and peripheral/autonomic neuropathy can be linked to AL amyloidosis ^6^. Frequent red flags that can be associated with AL amyloidosis onset or progression include gastrointestinal symptoms like nausea, vomiting, diarrhea, malabsorption, and weight loss (WL). As a result, the nutritional status of these patients can be compromised, and the cause is likely to be multifactorial ^7^ (figure 1).

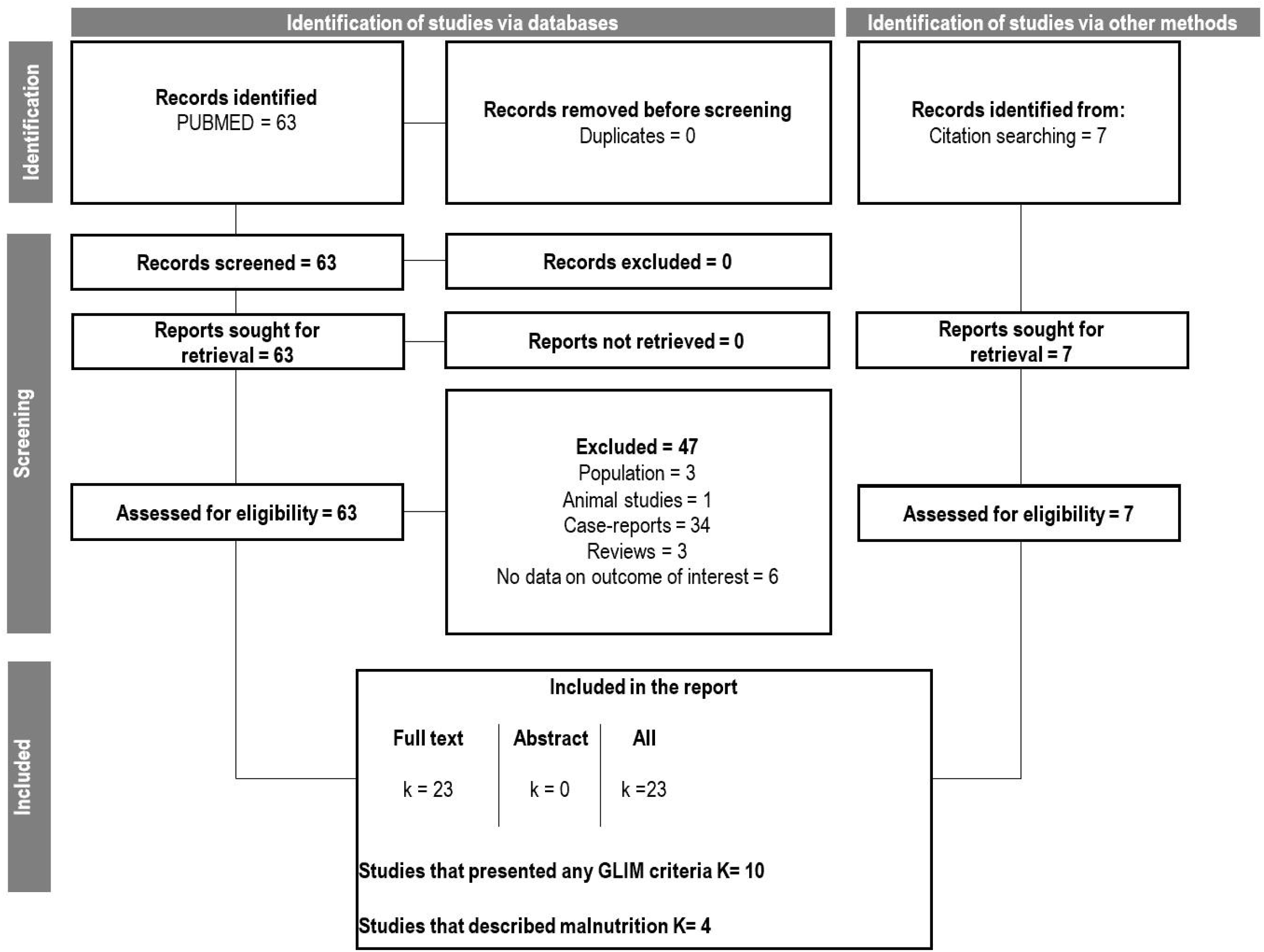

The nutritional status of patients is often neglected in the medical care pathway but may have considerable effects on physiological function ^8^. Poor nutritional status in patients with AL amyloidosis may be a consequence and/or be a factor that contributes to the progression of the disease. Malnutrition is known to have a negative impact on several outcomes that include reduced quality of life (QoL), increased morbidity and mortality, clinical complication rates, and length of hospital stay. All these elements contribute to the increasing burden of health costs ^9-11^. In this way, assessing nutritional status is an essential part of caring for patients with AL amyloidosis, to allow appropriate nutritional treatment and to prevent poor clinical outcomes. Several different criteria have been used to detect malnutrition in the general population. The most widely used are WL (with different cut points), anthropometry, body composition, reduced food intake, and subjective professional evaluation ^12-14^.

Although WL has been a symptom commonly reported in patients with AL amyloidosis ^6,15^, the nutritional status of these patients has not been deeply explored in the literature. A few studies have reported that malnutrition is highly prevalent and an independent predictor of survival among patients with AL amyloidosis ^16,17^. To date, there is no gold standard tool to assess nutritional status, neither does there exist a clear description of the clinical nutritional condition of these patients.

Our study aimed to review the available evidence on the nutritional status of patients with AL amyloidosis and its association with QoL and survival.

## METHODS

This rapid review was designed and conducted in accordance with the Preferred Reporting Items for Systematic Reviews and Meta-Analyses (PRISMA) guidelines ^18^.

### Data Sources and Search Strategy

The electronic search was conducted on PUBMED with a combination of MeSH terms and keywords searched for titles and abstracts. The following conjugated search terms were used: *(immunoglobulin light chain amyloidosis OR immunoglobulin light-chain amyloidoses OR amyloidosis, primary OR al amyloidosis OR al amyloidoses OR primary systemic amyloidosis OR amyloidoses, primary systemic OR amyloidosis, primary systemic OR primary systemic amyloidoses OR systemic amyloidosis, primary) AND (nutrition OR undernutrition OR malnutrition OR mal-nutrition OR malnourished OR underweight OR under-weight OR weight loss OR body mass index OR cachexia OR nutritional status)*.

The search was performed on August 12, 2020. There were no restrictions concerning the year of publication or language. Bibliographic references of the selected articles were manually screened for additional eligible studies by a single reviewer (SSG) and discussed with a second reviewer (PABR) as needed. We also contacted investigators by email to enquire about additional data when necessary. All retrieved data were screened using a bibliographic management program (EndNote®, version x9). The database was not blinded to the authors, institutions, or publication journals. All studies that provided enough information about the inclusion and exclusion criteria were selected for full-text assessment.

### Inclusion Criteria

Original cross-sectional, observational studies, cohorts and clinical trials were included. Studies on experimental animal models, reviews, and case-reports were excluded. In addition, studies that did not clearly report the nutritional data or for which the authors did not respond to our inquiry were also excluded.

## Data Extraction

Data extraction was performed by a single reviewer and revised by a second, using a pre-established data extraction form. Information extracted included author names, year of publication, country, study design, number of study participants, the age range, type of population, organ involvement, stage of the disease, and data of nutritional status (BMI, WL, malnutrition), in addition to outcomes of QoL and survival according to nutritional parameters.

### Data interpretation

Malnutrition was considered as defined by the authors. When lacking categorization of the outcomes of interest (i.e. BMI, WL and other nutritional parameters) they were extracted and presented according to the Global Leadership Initiative in Malnutrition (GLIM) ^19^. To diagnose malnutrition, based on GLIM criteria, at least one phenotypic criterion and one etiologic criterion should be present. The phenotypic criteria include: 1) non-intentional WL (>5% within past 6 months or >10% beyond past 6 months); 2) low BMI (<20 kg/m^2^ for adults <70 years, or <22 kg/m^2^ if 70 years and older); or 3) reduced muscle mass. The etiologic criteria include: 1) reduced food intake; 2) presence of gastrointestinal symptoms that cause malabsorption; and 3) disease-related inflammation.

## RESULTS

Our search strategy retrieved 62 records, and we identified 23 studies that satisfied the inclusion criteria (Figure 2 – Study flowchart). Table 1 describes the studies;; characteristics and outcomes of interest. The nutritional status of patients, such as malnutrition parameters, were often not clearly reported. For 10 studies, we were able to identify and extract at least one parameter recommended by GLIM criteria, even if partially fulfilled (details in Table 2). The cut points differed among studies, and none of them evaluated all the parameters necessary to completely fulfill the GLIM diagnosis criteria ^16,17,20,21,24,27,29,30,32,33^. Four studies presented a formal definition of malnutrition other than GLIM ^16, 17, 20, 32^.

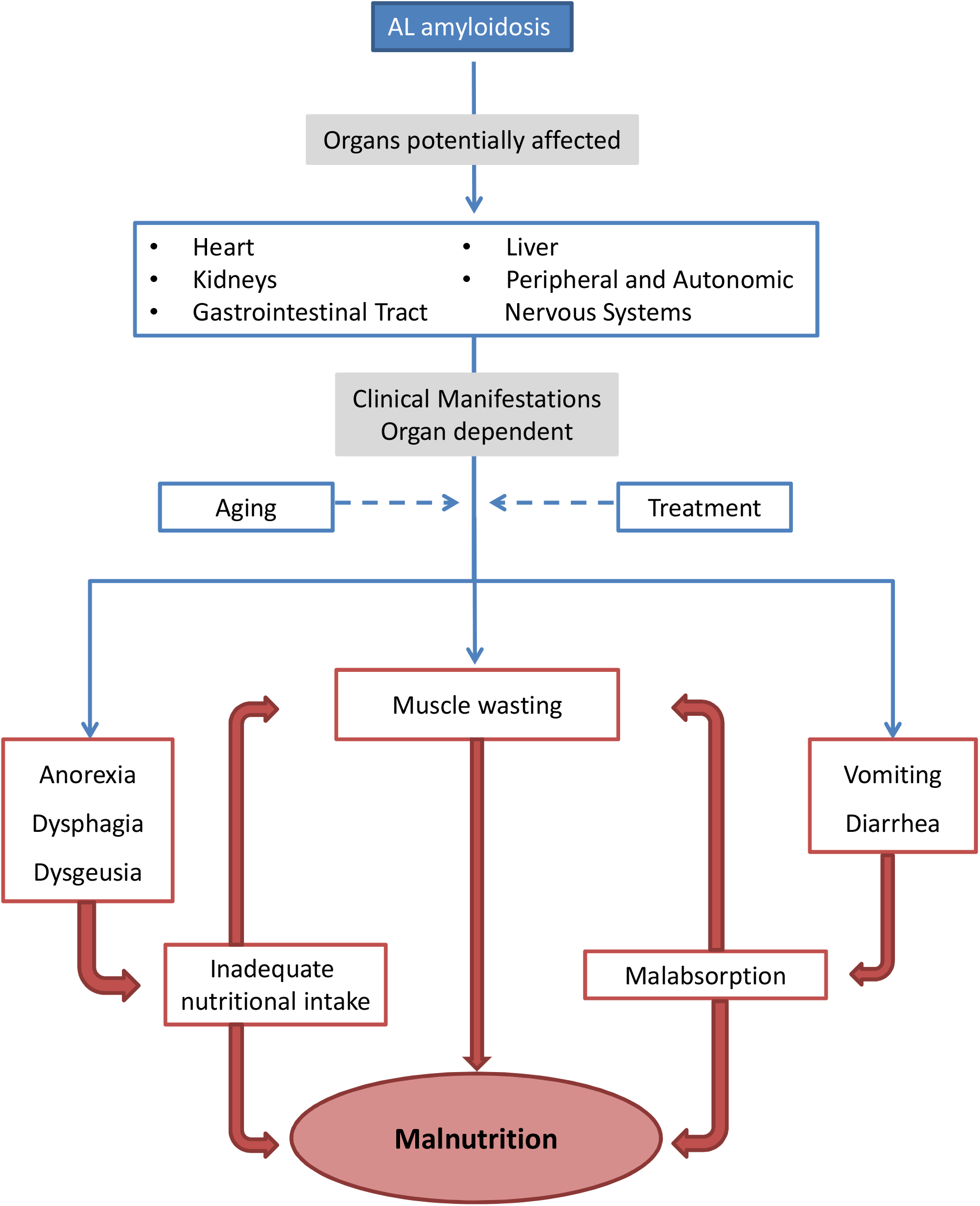

**Table 1.** Studies’ characteristics and summary of outcomes of interest according to nutritional status criteria.

| Author, Year; Country | Study population | Study design | Sample size; Age (years) mean ± SD; Sex proportions | Stage of disease; Organ/system involvement (%) | BMI (kg/m<sup>2</sup>) mean ± SD | Weight loss (Kg) mean ± SD | Symptoms GI (%) | QoL | Survival |
| --- | --- | --- | --- | --- | --- | --- | --- | --- | --- |
| Caccialanza R, 2020<sup>20</sup><br>Italy | Systemic AL Amyloidosis,<br>Newly diagnosed and treatment-naïve | Prospective cohort | n= 127<br>Age 64.1 ± 9.9<br>Male 54.3% | Mayo disease stage<br>I - 16.5<br>II - 33.1<br>IIIa - 23.6<br>IIIb - 26.8<br>Heart - 83.5<br>Kidney - 59.1<br>PNS - 8.7<br>Liver - 7.1 | 23.8 ± 3.7<br>C: <22<br>P: 32.3% | 8 ± 7.4<br>C: ≥10%<br>P: 31.5% | NR | PhA ≤4.3:<br>↓ PCS score (p=0.015)<br>MCS (p=0.16) | PhA ≤4.3:<br>HR 2.26<br>95%CI 1.04–4.89 |
| Bartier S, 2019<sup>21</sup><br>France | Cardiac amyloidosis (AL, ATTRwt, ATTRv), | Prospective cohort | AL n= 36<br>Age 67.66 ± 1.4<br>Male 66.7% | Prognostic stage<br>1 – 41.7<br>2 - 41.7<br>3 - 16.7<br>NR | median 22.9** (IQR 19.8–26.8)<br>C: <22<br>P: 41.7% | 6 ± 1.41**<br>P: 66.67%** | Dysphagia 25% | NR | NR |
| Szczygiel, JA, 2017<sup>22</sup><br>Poland | AL Cardiac amyloidosis<br>Newly diagnosed | Prospective cohort | n= 30<br>Age median 61.5 (IQR 39-75)<br>Male 53.3% | ECOG PS<br>1 – 23.3<br>2 – 43.3<br>3 – 10<br>4 – 23.3<br>NR | median 25.23 (IQR 23.5-27.9) | NR<br>C: >10kg<br>P: 70% | NR | NR | NR |
| Liewluck T, 2017<sup>23</sup>,<br>USA | Amyloid myopathy (isolated amyloid) | Cross-sectional | AL n= 32<br>Age median 65.5 | NR<br>Heart - 59.4<br>Kidney -15.6 | NR | NR<br>C: any WL<br>P: 28.13% | Dysphagia 33.25% | NR | NR |
| Mayo Clinic | myopathy and systemic) |  | (range 50-78.5)<br>Male 62.5% | GI - 12.5<br>Lung - 6.3<br>Liver - 3.1 |  |  |  |  |  |
| Decourt A, 2016<sup>24</sup>, France | End-stage renal disease AL, LCDD, or MCN | Retrospective cohort | AL n= 267<br>Age 70.6 ± 14.6<br>Male 64.4% | NR<br>NR | 23.2 ± 3.9<br>C: <20<br>P: 14% | NR | NR | NR | BMI <20:<br>HR 16.62<br>95%CI<br>11.21-23.61 |
| Caccialanza R, 2015<sup>7</sup>, Italy | AL Amyloidosis outpatients, treatment-naïve | RCT | n= 144<br>Age<br>NC: 65.4 ±10.5<br>UC: 64.8 ± 9.7<br>Male 61.1% | ECOG PS ≥2 - 55.6<br>Heart - 78.5<br>Kidney - 62.5<br>Liver - 13.2<br>GI - 4.9 | NC: 24.6 ± 3.3<br>UC: 25.4 ± 4.1 | NC: median 3.0 (IQR 0.0–6.0)<br>UC: median 4.0 (IQR 0.3–7.0) | Anorexia 45.9%<br>Dysgeusia 45.1%<br>Constipation 43.1%<br>Dysphagia 27%<br>Nausea 25.7%<br>Diarrhea 13.2%<br>Vomiting 6.9% | NC group:<br>↑MCS<br>8.1<br>95% CI<br>2.3–13.9<br>PCS 0.4<br>95% CI<br>-5.5 - 6.4 | NC group:<br>HR 0.57<br>95%CI<br>0.35–0.94 |
| Lim AY, 2015<sup>25</sup>, Korea | Systemic amyloidosis with gastrointestinal involvement (AL, AA, ATTR) | Retrospective cohort | AL n= 20<br>Age median 56 (range 37 - 72)<br>Male 55% | ECOG PS ≥2 - 35<br>Heart - 65<br>PNS - 31.6<br>Kidney - 30<br>ANS - 10.5 | NR | NR<br>C: any WL<br>P: 35% | Diarrhea 35%<br>Anorexia 30%<br>GI bleeding 25%<br>Nausea/vomiting 25%<br>Constipation 20%<br>Dyspepsia 15%<br>Reflux 15%<br>Abdominal pain 10% | NR | NR |
| Said SM, 2015<sup>26</sup>, USA<br>Mayo Clinic | Amyloidosis with gastrointestinal involvement (AA, AL, AApo A1, ATTR) | Retrospective cohort | AL n= 53<br>Age median 61 (range 43-96)<br>Male 64% | NR<br>Heart - 74<br>Kidney - 55<br>Nerve - 26<br>Liver - 15<br>Soft tissue/oral | NR | NR<br>C: any WL<br>P: 34% | Abdominal pain/dyspepsia 28%<br>Nausea/vomiting 23%<br>Dysphagia 23%<br>Diarrhea 8%<br>Early satiety 8%<br>GI bleeding 6% | NR | NR |
|  |  |  |  | cavity - 8 |  |  | Reflux 4% |  |  |
| Caccialanza R, 2014<sup>17</sup>, Italy | Systemic AL Amyloidosis, newly diagnosed and treatment-naïve | Prospective cohort | n= 128<br>Age 63.7 ± 9.9<br>Male 59.4% | ECOG PS ≥2 - 46.9<br>Heart - 78.1 | 25.2 ± 4.0<br>C: <22<br>P: 21.9% | median 3.0* (IQR 0.0-6.0)<br>C1: ≥10%<br>P1: 15.6% | NR | NR | BMI<22:<br>HR 1.98<br>95%CI<br>1.09 -3.56<br>WL*≥10%:<br>HR 1.94<br>95%CI<br>1.00 - 3.74<br>Moderate malnutrition:<br>HR 3.09<br>95%CI<br>1.75 - 5.46<br>Severe malnutrition:<br>HR 2.88<br>95%CI<br>1.23 - 6.72 |
| Caccialanza R, 2013<sup>27</sup>, Italy | AL amyloidosis, newly diagnosed and untreated | Prospective cohort | n= 187<br>Age 64.3 ± 9.5<br>Male 61% | NR<br>Heart - 79.1<br>Liver - 17.6 | 24.8 ± 3.8 | 5.5 (range 1.6–10)*<br>C: ≥10%*<br>P: 18.2% | NR | NR | NR |
| Sattianayagam PT, 2013<sup>16</sup>, UK | Systemic AL amyloidosis, newly diagnosed and treatment-naïve | Prospective cohort | n= 110<br>Age median 66 (range 42-88; IQR 60, 74)<br>Male 60% | Mayo disease stage<br>I- 14.5<br>II- 31.8<br>III- 53.7<br>Kidney - 77.3<br>Heart - 62.7<br>Liver - 14.6<br>ANS - 14.6 | median 25 (IQR 23 - 29) | NR<br>C: any WL<br>P: 60%* | Reduced appetite 43%<br>Dysgeusia 27%<br>Early satiety 25% | ↑ PG-SGA scores<br>↓ overall QoL (p<0.001) | Reference:<br>Score 0-1<br>PG-SGA score 4-8:<br>HR 6.2<br>95%CI<br>1.2 - 32.4<br>PG-SGA |
| | | | | PNS - 9.1<br>GI - 4.6 | | | | | score $\geq 9$:<br>HR 6.4<br>95%CI<br>1.4 - 30.2 |
| Cowan AJ,<br>2013<sup>28</sup>,<br>USA | Amyloidosis<br>with GI<br>involvement<br>(localized and<br>systemic AL,<br>ALys, ATTR,<br>ATTRw) | Retrospective<br>cohort | AL n= 50<br>Systemic<br>Age median<br>63 (range<br>46-79)<br>Male 64% | NR<br>Heart - 48<br>Nerves - 38<br>Kidney – 36 | NR | NR<br>C: any WL<br>P: 58% | Nausea 38%<br>Constipation<br>20%<br>Diarrhea 18%<br>Anorexia 14%<br>GI bleeding 8% | NR | NR |
| Hernández-Reyes J,<br>2012<sup>29</sup>,<br>Mexico | Systemic AL<br>amyloidosis | Retrospective<br>cohort | n = 23<br>Age median<br>57 (range<br>39-98)<br>Male<br>47.82% | NR<br>NR | NR | NR<br>C: >10%<br>P: 6% | NR | NR | NR |
| Caccialanza R,<br>2012<sup>30</sup>,<br>Italy | Systemic AL<br>amyloidosis,<br>newly<br>diagnosed | Prospective<br>cohort | n= 150<br>Age 65±8.9<br>(range 39–<br>87)<br>Males<br>61.3% | ECOG PS $\geq 3$ -<br>14<br>Heart - 83.3<br>Kidney - 63.3<br>Liver - 17.3<br>GI 10 - 6.7 | 24.6 ± 3.59<br>C: <22<br>P: 24.7% | median 5<br>(range 1 -<br>25)<br>P: 71.3%*<br>UBW<br>median<br>6.9%*<br>(range 1.2–<br>34.7)<br>C: $\geq 10\%$<br>P: 22%* | NR | WL> 10%<br>↓PCS<br>(p<0.001)<br>↓MCS<br>(p=0.003)<br>BMI< 22<br>↓PCS<br>(0.023)<br>MCS<br>(0.092)<br>MAMC<<br>10th<br>percentile<br>↓PCS<br>(0.007)<br>MCS (p=0.60) | NR |
| Madsen LG, 2009<sup>31</sup>, Denmark | AL amyloidosis - with gastrointestinal involvement | Retrospective cohort | n= 11<br>Age median 60 (range 50-69)<br>Male 63.6% | NR<br>Kidneys - 72.7<br>Heart - 54.5<br>Bone marrow - 54.5<br>Liver - 9<br>PNS - 36.7 | NR | median 7 (range 0-25)<br>C1: any WL<br>P1: 90.9%<br>C2: >10kg<br>P2: 36% | Diarrhea 45.5%<br>Reflux 36.4%<br>Constipation 9.1%<br>Abdominal pain 9.1%<br>GI bleeding 9.1%<br>Fecal fat 63.6% | NR | NR |
| Caccialanza R, 2006<sup>32</sup>, Italy | Systemic AL amyloidosis | Prospective cohort | n= 106<br>Age median 60 (range 28-78)<br>Male 56.6% | ECOG PS<br>0 -27.4<br>1- 45.3<br>2 - 17.9<br>3 - 9.4<br>Kidney - 74.5<br>Heart - 60<br>Liver - 24<br>GI - 2.8 | median 24.5 (rage 17.5-33.7)<br>C1: BMI<22<br>P1: 23.6%<br>C2: BMI<18.5<br>P2: 2.8% | median 8 (range 2-30)<br>UBW median 11.3% (range 2.6 - 34%)<br>C: any WL<br>P: 54.7% | Dysgeusia 34%<br>Anorexia 31.1%<br>Vomiting 17%<br>Dysphagia 14.1%<br>Diarrhea 8.5% | NR | BMI <22:<br>↓survival (P< 0.001) |
| Gertz MA, 2004<sup>33</sup> | Systemic AL amyloidosis | Pre and Post Multicenter Phase 2 study | N=28<br>Age median 54 (range 42-71)<br>Male 71% | NR<br>Kidney - 93<br>Liver - 25<br>Heart - 25<br>PNS - 18 | NR | NR<br>C1: <5%*<br>P1: 14%<br>C2: 5-10%*<br>P2: 4%<br>C3: >10%*<br>P3: 7% | NR | NR | NR |
| Park, MA, 2003<sup>34</sup>, USA<br>Mayo Clinic | Systemic AL amyloidosis with liver involvement | Retrospective cohort | n= 98<br>Age median 58.5<br>Male 69% | NR<br>Kidney - 33<br>Heart - 12<br>PNS - 6 | NR | 10.4<br>C: any WL<br>P: 72% | Abdominal pain 53%<br>Anorexia 26%<br>Early satiety 19%<br>Nausea 15%<br>Dysgeusia 10% | NR | NR |
| Hayman SR, 2001<sup>35</sup>, USA<br>Mayo Clinic | Systemic AL amyloidosis with malabsorption syndrome | Retrospective cohort | n= 19<br>Age median 63 (range 41-74)<br>Male 74% | NR<br>Heart - 77<br>Bone marrow - 36 | NR | median 30 (range 2-134)<br>C1: any WL<br>P1: 100%<br>C2: >20lb<br>P2: 68.4% | Diarrhea 68.4%<br>Anorexia 42.1%<br>Steatorrhea, nausea, or Abdominal pain 26.31%<br>Vomiting or edema 21% | NR | WL ≤20 lb: median survival - 22 months<br>WL >20 lb: median survival - 10 months |
| Dispenzieri A, 2001<sup>36</sup>, USA<br>Mayo Clinic | Systemic AL amyloidosis | Retrospective cohort | n= 229<br>Age median 56 (range 25 - 70)<br>Male 58% | ECOG PS<br>0 - 40<br>1 - 41<br>2 - 12<br>3 - 5<br>Kidney - 73<br>Heart - 44<br>Liver - 18<br>Nerve - 17<br>GI - 3.9<br>Lung - 1.31 | NR | NR<br>C1: ≤10lb<br>P1: 60.3%<br>C2: >10lb<br>P2: 39.7% | NR | NR | WL > 10 lb<br>↓survival<br>RR 1.5<br>95% CI 1.1-2.1 |
| Dubrey SW, 1998<sup>37</sup>, USA | AL amyloidosis with heart involvement | Retrospective cohort | n= 232<br>Age 60 ±11 median 59 (range 29–85)<br>Male 61% | NR<br>Heart - 100<br>GI - 68<br>Kidney - 67<br>Nervous System - 55<br>Musculo- | NR | NR<br>C1: ≥10lb<br>P1: 42.7%<br>C2: 10-20lb<br>P2: 18.1% | Early satiety/ anorexia 22.8%<br>Nausea 22.8%<br>Dysphagia 6%<br>Diarrhea 29.3%<br>Dysgeusia 10.3%<br>Constipation 6.0% | NR | NR |
|  |  |  |  | Skeletal - 31<br>Skin - 25<br>Lung - 16 |  | C3: 21-30lb<br>P3: 12.9%<br>C4: 31-40lb<br>P4: 4.3%<br>C5: >50lb<br>P5: 7.3% | Abdominal<br>discomfort<br>10.8% |  |  |
| Kyle RA,<br>1995<sup>38</sup><br>USA<br>Mayo Clinic | Systemic AL<br>amyloidosis | Retrospective<br>cohort | n= 474<br>Age median<br>64 (range<br>32-90)<br>Male 69% | NR<br>Kidney - 30<br>Heart - 22<br>PNS - 17 | NR | median 23lb<br>(range 4-200)<br>C: any WL<br>P: 52% | NR | NR | NR |
| Menke, DM,<br>1993<sup>39</sup>,<br>USA<br>Mayo Clinic | Systemic AL<br>amyloidosis<br>with<br>Symptomatic<br>Gastric<br>Amyloidosis | Retrospective<br>cohort | n= 8<br>NR<br>NR | NR<br>NR | NR | (range 6.5-22.5)<br>C: any WL<br>P: 87.5% | Hematemesis or<br>prolonged<br>nausea and<br>vomiting 100% | NR | NR |

\* Within the past 6 months; \*\* data provided by the author; BMI - body mass index: values are mean ± standard deviation; WL - weight loss: values are mean ± standard deviation; MAMC - mid-arm muscle circumference; QoL - Quality of life; SD – standard deviations; IQR - interquartile range; C - criteria; P - prevalence; NR – not reported; ECOG PS – Eastern Cooperative Oncology Group performance status; GI - gastrointestinal; MCS - mental component summary; NC - nutritional counseling; PG-SGA score – Patient-Generated Subjective Global Assessment; PNS - peripheral nervous system; UBW - usual body weight; UC usual care; AA - Secondary amyloidosis (associated with deposition of serum amyloid A protein); AApo A1 - apolipoprotein A1 amyloidosis; AL - immunoglobulin light chain amyloidosis; ALys - Hereditary lysozyme (ALys) amyloidosis; ANS - Autonomic nervous system; ATTR- amyloidosis transthyretin-related; ATTRwt - transthyretin wild-type; ATTRv – transthyretin mutant form; LCDD light – chain deposition disease; MAMC - mid-arm muscle circumference; MCN - myeloma cast nephropathy;

**Table 2.** Cut-off point variation of malnutrition parameters, and agreement with GLIM criteria.

|  | Phenotypic parameters |  |  | Etiologic parameters |  |  | GLIM criteria agreement (3/3) |
| --- | --- | --- | --- | --- | --- | --- | --- |
|  | Weight Loss >10%<br>% of patients<br>(time frame) | BMI<br>% of patients<br>cut-off point | Low muscle mass<br>% of patients<br>Measure; cut-off point | Reduced food intake or assimilation |  | Inflammation<br>% of patients;<br>criteria | Phenotypic (n)/<br>Etiologic (n) |
|  |  |  |  | % of Estimate Energy Requirements<br>% of patients<br>cut-off point | Gastrointestinal condition<br>% patients;<br>condition |  |  |
| Caccialanza R (2020)<sup>20</sup> | 31.5% (6 months)* | 32.3%<br><22Kg/m<sup>2</sup> | NR | 76.2%<br><75% of estimate | NR | 100% chronic disease* | 1/1 |
| Bartier S (2019)<sup>21</sup> | NR | 41.7%<br><22Kg/m<sup>2</sup> | NR | NR | NR | 100% chronic disease* | 0/1 |
| Decourt A (2016)<sup>24</sup> | NR | 14%<br><20Kg/m<sup>2</sup> | NR | NR | NR | 100% chronic disease* | 0/1 |
| Caccialanza R (2014)<sup>17</sup> | 15.6% (6 months)* | 21.9%<br><22Kg/m<sup>2</sup> | NR | NR | NR | 100% chronic disease* | 1/1 |
| Caccialanza R (2013)<sup>27</sup> | 18.2% (6 months)* | NR | NR | NR | NR | 100% chronic disease* | 1/1 |
| Sattianayagam PT (2013)<sup>16</sup> | NR | <4%<br><20Kg/m<sup>2</sup> | NR | NR | 4.6%*<br>Gastrointestinal involvement | 100% chronic disease* | 0/2 |
| Hernández-Reyes J (2012)<sup>29</sup> | 6% (NR) | NR | NR | NR |  | 100% chronic disease* | 0/1 |
| Caccialanza R | 22% (6 months)* | 24.7% | 45.3%* | 24.6% | 6.7%* | 100% chronic | 2/2 |
| (2012)<sup>30</sup> |  | <22Kg/m<sup>2</sup> | MAMC <10<sup>th</sup> percentile | <60% of estimate | Gastrointestinal involvement | disease* |  |
| Caccialanza R (2006)<sup>32</sup> | NR | 23.6% <22Kg/m<sup>2</sup><br>2.8% <18.5Kg/m<sup>2</sup> | 27.3%*<br>MAMC <10<sup>th</sup> percentile | NR | 34% Dysgeusia<br>31.1% Anorexia<br>17% Vomiting<br>14.1% Dysphagia<br>8.5% Diarrhea* | 100% chronic disease* | 1/2 |
| Gertz MA (2004)<sup>33</sup> | 7% (6 months)* | NR | NR | NR | NR | 100% chronic disease* | 1/1 |

NR: Not Reported. \*Assessment parameter and cut-off point completely in accordance with GLIM criteria.

### Characteristics of the studies

Studies were published between 1993 and 2020. Most of the studies were conducted in North America (10: USA ^23, 26, 28,33-39^), and Europe (11: England ^16^, France ^21, 24^, Italy ^7, 17, 20, 27, 30, 32^, Poland ^22^, Denmark ^31^). One was from Asia (Korea ^25^), and one from Central America (Mexico ^29^). Eight studies from the USA are from the same database (Mayo Clinic) ^23, 26, 33-36, 38, 39^, and the 6 Italian studies are from the same institution ^7, 17, 20, 27, 30, 32^.

In these studies, sample sizes ranged from 8 to 474 patients, totaling 2562 patients, though there is likely overlap of patients across studies from the center^7, 17, 20, 23, 26, 27, 30, 32, 33-36, 38,39^. Overall, men accounted for 61.7% of the population, age ranged from 25 to 98 years. The studies included patients with various principal organ(s) involvement and different stages of the disease. Cohort studies were predominant, and only two studies presented data from interventional trials: one study had a randomized controlled clinical trial design ^20^, and another was a phase 2 clinical trial ^33^.

### Malnutrition

Four studies ^16, 17, 20, 32^ presented a formal malnutrition diagnosis according to four different criteria, with a range of prevalence from 25% to 65%. Caccialanza et al. ^20^ reported that 52% of the AL amyloidosis patients had malnutrition according to the phase angle (PhA) derived from bioelectrical impedance. The authors used the median value of the sample as the cut point to detect malnutrition (PhA ≤ 4.30□). One study ^16^ assessed malnutrition using the Patient-Generated Subjective Global Assessment (PG-SGA) score. This study showed that 65% of the patients had a PG-SGA score equal to or greater than 4 (defined as malnutrition by the authors). No correlation between BMI and nutritional status by PG-SGA was found (r=-0.14). Cacciallanza et al. ^17^, assessed malnutrition according to BMI < 22 kg/m^2^ and unintentional WL in the previous 6 months (WL when ≥ 10%). The results showed a prevalence of moderate (presence of one parameter) and severe malnutrition (both parameters combined) of 21.9% and 7.8%, respectively. Cacciallanza et al. ^30^ considered a cut point of BMI < 22 kg/m^2^ as a criterion for malnutrition and classified nearly 25% of the patients as malnourished. They also reported BMI < 18.5 kg/m^2^, a classification of underweight according to the WHO, in 2.8% of patients ^40^.

### Weight Loss

Detailed data of WL is presented in Table 1. Of the 23 selected studies, only one ^24^ did not provide any data on WL. Twelve ^7, 17, 20, 21, 27, 30^-^32, 34, 35, 38, 39^ studies presented WL as descriptive data (mean, median or range) ranging from 0 to 30 kg or from 2 to 200lb. Fourteen studies highlighted the prevalence of any amount of WL, and it ranged from 28.3% to 100%^16,21,22,25,26,28,30-35,38,39^. Considering a percentage of WL greater than 10%, six studies reported a prevalence ranging from 6% to 31.5% of the patients ^17,20,27,29,30,33^. The results from the RCT demonstrated that an intervention with nutritional counseling during 12 months allowed patients with systemic AL amyloidosis to maintain their weight (WL = 0.6 kg; *p* = 0.214), while patients in the usual care group had a significant decrease (WL = 2.1 kg; *p* = 0.003) ^20^.

### Body Mass Index

Table 1 presents the studies that described data of BMI in patients with AL amyloidosis (k=10). All of them reported mean (±SD) or median (range/IQR) BMI. The mean ranged from 19.3 to 29.2 kg/m^2^, and the median ranged from 17.5 to 33.7 kg/m^2^ ^7, 16, 17, 20^-^22, 24, 27, 30, 32^. Seven studies presented BMI with different cut points. The percentage of patients with a BMI < 22kg/m^2 17,20,21,30,32^ ranged from 21.9% to 41.7% and from 3.63% - 14% for BMI <20kg/m^2^ ^16, 24^.

### Association between self-reported quality of life and nutritional status

Four studies ^7, 16, 20, 30^ investigated the association between nutritional status and QoL, using the Medical Outcomes Study 36-item Short-Form General Health Survey (SF-36), ^7, 20, 30^ or the European Organization for Research and Treatment of Cancer (EORTC) QLQ-C30 ^16^. Sattianayagam et al. ^16^, demonstrated that more severe malnutrition evaluated by the PG-SGA score correlated with lower QoL scores (*p*< 0.001), including lower functional and higher symptom scores. Calccialanza et al. ^30^, demonstrated that the mental component summary was decreased by 0.47 (95%CI 0.18-0.75, *p*= 0.002) for each kilogram of WL in the previous 6 months. In addition, one study observed that patients with a lower value of the PhA had lower Physical Health Composite Scale (PCS) scores (*p*=0.015)^20^. Finally, the randomized clinical trial showed a significant increase in the mental component summary after receiving nutritional counselling, and patients were able to achieve a mean score of 53 (95% CI, 50–53), which is above the healthy population norms ^7^.

### Association between survival and nutritional parameters

Overall, 7 studies reported survival rates according to nutritional parameters (details in Table 1) ^16,17,20,24,32,35,36^. The results were presented as hazard ratios or relative risk, with a range from 0.75 to 6.4, depending on the parameter and cut point analyzed. A score higher than 9 on PG-SGA was associated with higher mortality (HR = 6.4; 95%IC 1.4-30.2)^16^. Weight loss, BMI < 22kg/m2 and low PhA (≤ 4.30□) were also independently associated with mortality ^17,20,32,36^.Furthermore, patients who received nutritional counseling had a lower mortality risk than those who did not (HR 0.57; 95%IC 0.35-0.94)^7^.

## DISCUSSION

Our results suggest that AL Amyloidosis has a negative impact on the nutritional status of patients. Most of the evidence on nutritional status is derived from retrospective data. The included studies provided nutritional status prevalence that varied markedly depending on the assessment tool, criteria used, organs involved, and disease stage. The prevalence of malnutrition in AL amyloidosis patients ranged from 25% to 65%. This wide range is likely a consequence of inconsistent diagnostic criteria across studies and the disease’s heterogeneity, but nevertheless the results indicate that malnutrition is consistently highly prevalent.

Caccialanza et al. ^20^, using BIA technology, found that 52% of patients had malnutrition. While this measurement technique is objective, easy to perform, non-invasive and inexpensive^53^, the cut point to detect malnutrition is unclear. The authors used the median of the PhA as a diagnostic measure and as a prognostic threshold value. Low values of PhA have been previously associated with malnutrition according to different cut-off points varying from 4.0° to 6.0°^54^. One study assessed malnutrition with PG-SGA^16^ and showed the highest prevalence of malnutrition (65% of the patients), probably because this tool uses a combination of parameters (i.e., WL, dietary intake, gastrointestinal symptoms, functional capacity, nutritional requirements according to the disease and physical evaluation). In the absence of a gold standard method to detect malnutrition, the SGA is considered one of the best assessment tools since it has been validated and is predictive of outcomes in many clinical conditions, including cancer ^41-43^. In the studies of Caccialanza et al. ^7, 27, 30, 32^, mid-arm muscle circumference, caloric and protein intake were assessed in combination with BMI and WL to better evaluate the nutritional status.

These findings are consistent with those that showed malnutrition in patients with similar characteristics of our studied population. Despite also having a large variance in diagnosis and reporting, malnutrition has a prevalence ranging from 16% to 62.4% in stable heart failure ^44^, and it is as high as 80% in patients with advanced heart failure^45^. Among older cancer patients under chemotherapy, malnutrition studies showed prevalence from 3 to 83% ^46^. Also, aging itself may also induce deterioration in nutritional status due to multiple factors ^47^, which could partially explain the high prevalence we observed.

The highest inconsistency was found in reporting the prevalence of WL, that ranged from 6% to 100% due multiple cut-off points (e.g. WL> 10kg ^22, 31^, WL ≥10 lb ^36, 37^, WL > 20 lb ^35, 37^, WL >100 lb ^38^, WL≥ 5% and WL≥ 10% of the usual body weight ^17, 20, 27, 29, 30, 33^). Interestingly, most studies did not even use a cut point and presented the prevalence based on any amount of WL ^16,21,23,25,26,28,30-35,38,39^. Moreover, most studies have not indicated the timeframe for WL, which is critically relevant. These inconsistencies render it difficult to interpret and compare the data across studies. Furthermore, many patients with systemic AL amyloidosis experience fluid retention, usually due to cardiac and renal involvement. This can impair accurate assessment of nutritional status by WL and BMI ^48^ and also interfere with the interpretation of the results.

According to BMI, the prevalence of malnutrition in this population ranged from 2.8% to 41.7% ^16,17,21,24,30,32^. Although low BMI is commonly accepted in the medical community, used alone it is a poor indicator of malnutrition, and can misclassify malnourished patients within the normal range ^49^. Perhaps, for this reason, the study that assessed malnutrition using this criterion alone had the lowest prevalence. Also, BMI may not be reliable in the presence of confounding factors, such as edema or ascites, and may not identify significant unintended WL when used as a single assessment^49^. Therefore, we believe BMI should be used in combination to WL and/or other parameters to better evaluate malnutrition.

Quality of life among patients with AL Amyloidosis has not been extensively studied^16, 30, 50-52^. However, some studies have already shown that the impact of the disease results in broad health-related QoL deficits across physical and mental domains when compared to the general population^30,50-52^. Overall, QoL seems to be associated with nutritional status ^16, 20, 30^ independently of other relevant clinical variables, including disease stage, organ involvement and age^30^. Interestingly, the only randomized clinical trial with nutritional counseling intervention showed an improvement in the mental component of QoL, but no effect on physical component^7^. There was also a trend toward maintaining body weight, improve calorie intake, and survival ^7^.

Survival in AL amyloidosis depends greatly on the stage of the disease and the organs in which light chain fibrils are deposited. Poor nutritional status, however, despite being a prevalent condition among these patients, has thus far garnered little attention in survival analyses. Furthermore, although malnutrition is a public health issue already proven to be associated with incremental morbidity, mortality, and health costs, there has been a lack of evidence in particular settings^19^, including amyloidosis patients. In our review, 7 studies tested the association between nutritional status and survival^16, 17, 20, 24, 32, 35, 36^ all using different nutritional parameters, making it difficult to determine which assessment tool should be used and the real impact on survival.

Finally, assuming that nutritional status is a modifiable risk factor, the evidence from our review reinforces the importance of a standardized nutritional status assessment, enabling tailored nutritional intervention as soon as possible. Patients with AL amyloidosis have a broad spectrum of symptoms, and for that reason we believe that diagnostic criteria covering different domains, instead of using single parameters, would be more appropriate. In this context, the GLIM consensus is an initiative led by the American Society for Parenteral and Enteral Nutrition (ASPEN) proposing criteria that consider a combination of phenotypic and etiologic parameters. In light of the complexity of the clinical manifestations, we strongly believe that a more holistic approach such GLIM could successfully be used in clinical practice and optimize patients’ management.

Some limitations require acknowledgment in this review. First, we considered only PubMed retrievals; using the identical search strategy on EMBASE yielded zero retrievals. Second, most of the studies mentioned did not report nutritional status as a primary outcome, which underscores the need for more data. Finally, due to the different criteria and cut points used to assess malnutrition and WL, quantitative summarization of the results was not possible (i.e., meta-analysis).

## CONCLUSION

In conclusion, our review shows a high prevalence of WL and malnutrition in AL amyloidosis patients. Our study also has many strengths, such as methodological rigor and being the first to analyze the evidence and highlight the importance of properly assess nutritional status and its potential impact on patients’ QoL and survival. As most of the evidence comes from retrospective studies, it is difficult to evaluate the disease’s true impact on nutritional status and its association with QoL and survival. We believe that criteria standardization and proper assessment tools will promote better understanding. It is clear that some effort should be invested in testing and validating accurate tools in this population, such as the GLIM criteria. Overall, the available data suggest that we need to improve assessment to be able to offer treatment to optimize health care in this population.

## Data Availability

The data used to support the findings of this study are included within the articles

## Acknowledgments

We thank Dr. Annabel Chen-Tournoux for the critical review of this manuscript.

## Authors’ contributions

Conception and design of the research: SSG, PZ, PABR, FT; Analysis and interpretation of the data: SSG, PZ, PABR, FT; Writing of the manuscript: SSG, PZ, PABR, FT; Critical revision of the manuscript for intellectual content: PABR, FT; Final review: SSG, SB, PABR, ELB, BL, FT.

### Funding support

Scholarship from Fonds de recherche du Quebec - Nature et Technologies (FRQNT) for SSG. FT is supported by a grant from Fonds de recherche du Québec – Santé. This research did not receive any other specific grant from funding agencies in the public, commercial, or not-for-profit sectors.

